# Diagnostic Accuracy in Bronchial Carcinoid tumors is Dependent of Biopsy Size

**DOI:** 10.1101/2021.05.20.21257521

**Authors:** E.M.B.P. Reuling, D.D. Naves, J.M.A. Daniels, C. Dickhoff, P.C. Kortman, M.A.M.B. Broeckaert, P.W. Plaisier, Erik Thunnissen, T. Radonic

**Affiliations:** Department of Surgery, Amsterdam University Medical Center, VU University Amsterdam, De Boelelaan 1117, 1081 HV Amsterdam, the Netherlands; Department of Cardiothoracic Surgery, Amsterdam University Medical Center, VU University Amsterdam, De Boelelaan 1117, 1081 HV Amsterdam, the Netherlands; Department of Pathology, Amsterdam University Medical Center, VU University Amsterdam, De Boelelaan 1117, 1081 HV Amsterdam, the Netherlands; Department of Pulmonary Diseases, Amsterdam University Medical Center, VU University Amsterdam, De Boelelaan 1117, 1081 HV Amsterdam, the Netherlands; Department of Cancer Centre Amsterdam; De Boelelaan 1117, 1081 HV Amsterdam, the Netherlands; Department of Surgery, Albert Schweitzer Hospital, Albert Schweitzerplaats 25, 3318 AT Dordrecht, the Netherlands

**Keywords:** Typical carcinoid, Atypical carcinoid, Biopsy size, Endobronchial therapy

## Abstract

**Objective:** Recently, 60% discordancy was reported for distinction between typical carcinoid and atypical carcinoid in preoperative biopsy compared to the resection specimen. This study investigated the impact of biopsy surface size, obtained with flexible and rigid bronchoscopy, on diagnostic accuracy of typical and atypical carcinoid.

**Methods:** Biopsy-resection paired specimens of patients referred for treatment to Amsterdam University Medical Centers were retrieved. Bronchial biopsies were obtained either by flexible or rigid biopsy. The definitive diagnosis was based on the resection specimen. Diagnosis according to the 2015 WHO classification, mitoses and necrosis in biopsy and resection specimen, were independently re-evaluated by two pathologists.

**Results:** After screening 298 patients, 64 biopsy-resection pairs with available tissue were included of which 34 (53%) were biopsied with flexible and 30 (47%) with rigid biopsy. In 35 (55%) patients, the tumor classification between the biopsy and resection specimen was concordant. The discordance in the remaining 29 cases (45%) was caused by misclassification of atypical as typical carcinoid in bronchoscopy specimens, predominantly in small flexible biopsies (59%, *p*=0.021). Of biopsies measuring <2 mm^2^, 79% were classified as discordant and 52% of the discordant biopsies measured <4 mm^2^.

**Conclusion:** Histological classification in central carcinoid tumors is discordant in 45% of the biopsies, with increasing diagnostic accuracy in larger biopsies. Distinguishing carcinoid tumor into typical or atypical carcinoid on biopsies <4 mm^2^ should be discouraged. A cumulative biopsy surface of at least 4 mm^2^ tumor is preferred to increase the diagnostic accuracy which helps in optimal treatment planning.

## Introduction

Pulmonary carcinoids comprise a subgroup of neuroendocrine tumors and are categorized into low-grade typical carcinoid (TC) and intermediate-grade atypical carcinoid (AC) according to the current WHO classification (WD Travis, 2015). Morphologically, TC is defined as a neuroendocrine tumour with less than 2 mitoses per 2 mm^2^ and absence of necrosis, while AC has 2-10 mitoses per 2 mm^2^ and/or dot-like necrosis (WD Travis, 2015, Travis, 2014). Accurate identification of carcinoid tumors and morphologic distinction between central AC and TC at time of diagnosis is important as it directs treatment selection. For example, endobronchial treatment is a promising parenchyma sparing procedure for selected patients with centrally growing intraluminal bronchial TC. During this parenchyma-sparing procedure, a rigid bronchoscope is used which allows for larger biopsies and in selected cases even complete resection (Reuling et al., 2018). Furthermore, diagnostic accuracy might implicate a more aggressive search for potential dissemination as AC tend to metastasize more often than AC (Travis, 2014). It was recently showed that classification of carcinoids based on pre-operative biopsies is imprecise (Moonen et al., 2020), resulting in 57% discordance when compared with definitive, postoperative pathology. In the current study, we hypothesized that concordance between pre- and postoperative pathology improves with larger preoperative biopsies. To test this hypothesis, we investigated the relation of biopsy surface accuracy of diagnosing TC and AC correctly.

## Material and methods

Approval of the institutional review board (Medical Ethics Review Committee of VU University Medical Center, IRB00002991) and consent from each patient after full explanation of the purpose of this study was retrieved. Patients who underwent surgical resection for centrally located pulmonary carcinoid (stage I-III) between June 1991 and December 2019 at the Amsterdam University Medical Center, were screened for eligibility. Central tumors where defined as tumors situated proximal to the segmental bronchi. Patients who had paired diagnostic biopsies obtained with either Flexible (FLB) or rigid (RIB) biopsy, were selected. Samples of central carcinoid tumors were independently re-evaluated by two pathologists (TR & ET), and scored for mitotic count, presence of necrosis and diagnosis. Tumor classification on the resection specimen was considered as the gold standard, and thus marked as definitive diagnosis. Mitotic figures on biopsies and resection specimen were counted as described previously (Baak, 1991). In short, the whole slide was first explored for mitotic hotspots and the mitotic count was subsequently performed in the hotspot area. HE stained slides were scanned using Phillips UFS scanner and analyzed with the Philips pathology viewer version 3.2. Histological tumor sample size was digitally measured and defined as tumor surface (mm^2^) in the whole histological sample. Areas with cauterization or mechanical artifacts were discarded. The statistical analyses and calculations were performed with IBM SPSS Statistics for Windows, version 26 (IBM Corp., Armonk, N.Y., USA) (ER, DN).

## Results

After screening 298 patients, paired biopsy and resection specimens of central pulmonary carcinoids from 64 patients were available. The diagnosis was based solely on mitotic count, as (dot-like) necrosis was absent. No significant differences were observed between patients with TC (*n*=26) and AC (*n*=38) regarding clinic-pathological characteristics, except for a trend of a larger tumor diameter in AC patients (*p*=0.05) (Table 1).

**Table 1:** Clinicopathological characteristics of central carcinoid cohort.

| Patient characteristics | N=64 (%) | TC (n=26) | AC (n=38) | p-value |
| --- | --- | --- | --- | --- |
| **Mean age in years (SD)** | 45 (16.2) | 43 (15.8) | 46 (16.7) | 0.51 |
| **Female** | 40 (62.5) | 14 (53.8) | 25 (65.8) | 0.33 |
| **Biopsy** |  |  |  |  |
| FLB | 34 (53.1%) | 12 (46.2%) | 22 (57.9%) | 0.36 |
| RIB | 30 (46.9%) | 14 (53.8%) | 16 (42.1%) |  |
| **Type of surgery** | 64 | 26 | 38 | 0.10 |
| Pneumonectomy | 3 (4.7) | 2 (7.7) | 1 (2.6) |  |
| Bilobectomy | 13 (20.3) | 9 (34.6) | 5 (13.2) |  |
| Lobectomy | 29 (45.2) | 8 (30.7) | 20 (52.7) |  |
| Sleeve lobectomy | 17 (26.6) | 6 (23.1) | 11 (28.9) |  |
| Segmentectomy | 1 (1.6) | 1 (3.8) | 0 |  |
| Bronchial sleeve resection | 1 (1.6) | 0 | 1 (2.6) |  |
| **Tumor diameter** |  |  |  | 0.05 |
| T1a | 38 (59.4) | 21 (80.8) | 17 (44.7) |  |
| T1b | 14 (22) | 3 (11.5) | 11 (28.9) |  |
| T2a | 8 (12.5) | 2 (7.7) | 6 (15.8) |  |
| T2b | 2 (3.1) | 0 | 2 (5.3) |  |
| T3 | 2 (3.1) | 0 | 2 (5.3) |  |
| T4 | 0 | 0 | 0 |  |
| **Nodal stage** |  |  |  | 0.78 |
| N0 | 59 (92.2) | 25 (96.2) | 34 (89.5) |  |
| N1 | 4 (6.3) | 1 (3.8) | 3 (7.9) |  |
| N2 | 1 (1.6) | 0 | 1 (2.6) |  |
| **Radicality** |  |  |  | 0.68 |
| R0 | 58 (90.6) | 23 (88.5) | 35 (92.1) |  |
| R1 | 6 (9.4) | 3 (11.5) | 3 (7.9) |  |

Figure 1 presents the distribution of diagnoses (TC vs AC), mitotic count and histological tumor sample size for flexible biopsy, rigid biopsy and surgical resection specimen respectively. For all biopsies, the diagnosis TC or AC was concordant with definitive pathology in 35 out of 64 patients (55%). In the remaining 29 (45%) patients, the biopsy-based diagnosis was TC while the diagnosis in the pulmonary resection specimen was AC. When considering biopsies obtained with FLB and RIB separately, discordance was 59% and 30%, respectively (*p*=0.021). In total, 38 (59%) cases were identified with definitive AC diagnosis. Nine histological AC diagnoses were made in the biopsies, more often in RIB (7/30, 23%) than in FLB (2/34, 6%) (*p*=0.07, Fig. 1A and Table 2). In RIB, a higher number of mitotic figures and a larger histological tumor sample size were demonstrated when compared with FLB (*p*=0.012 and *p*<0.001, respectively; Fig. 1B & 1C).

**Table 2:** accuracy between diagnosis of flexible (FLB) and rigid (RIB) biopsy and resection. Discrepancies are underscored.

|  | Diagnosis resection |  |  |
| --- | --- | --- | --- |
|  | TC | AC | Total |
| Diagnosis FLB TC | 12 | <u>20</u> | 32 |
| Diagnosis FLB AC | 0 | 2 | 2 |
| Diagnosis RIB TC | 14 | <u>9</u> | 23 |
| Diagnosis RIB AC | 0 | 7 | 7 |

**Figure 1:**
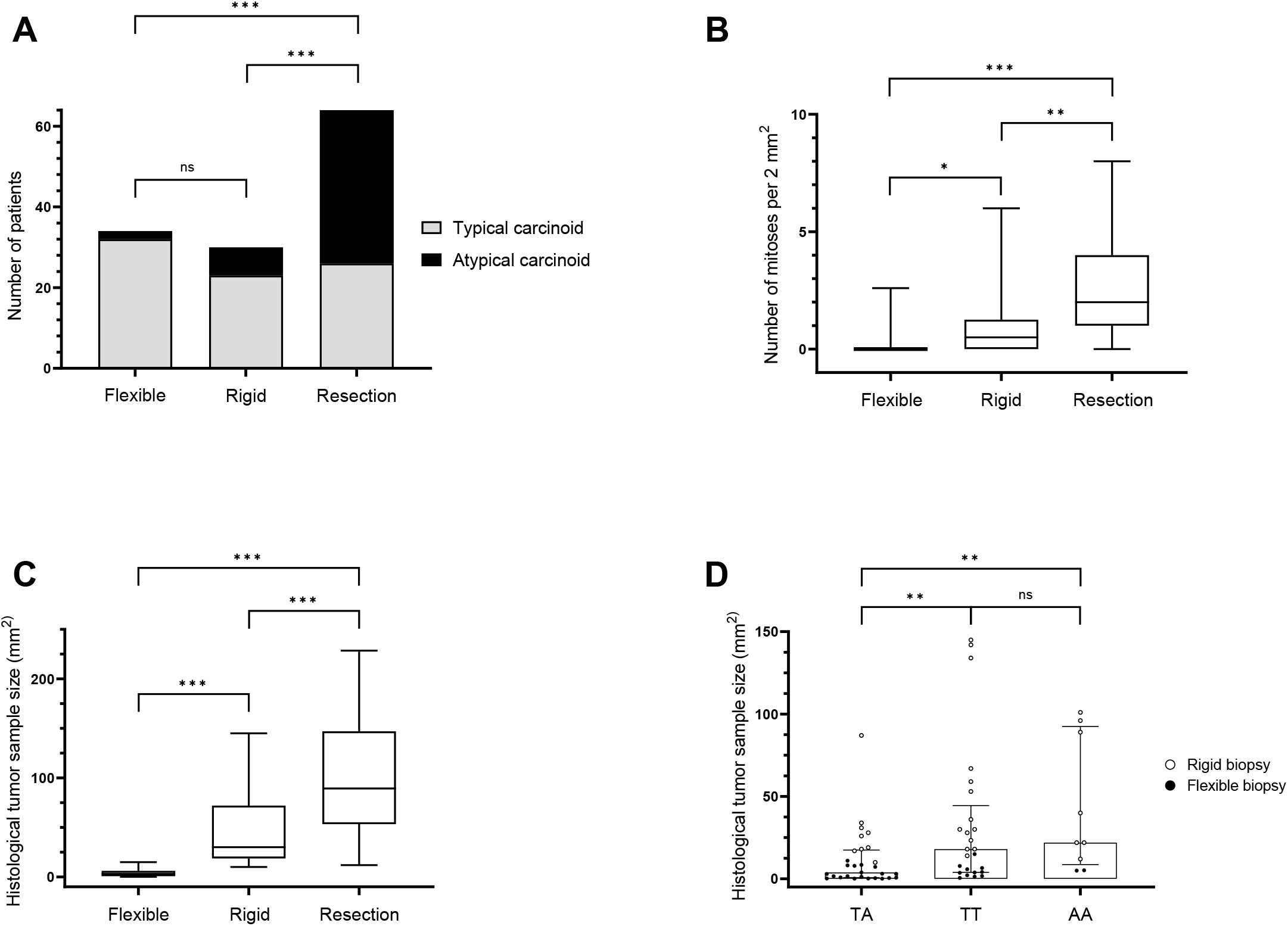
Outcomes in flexible biopsy, rigid biopsy and resection in relation to typical and atypical carcinoid (A), mitotic count (B), and surface area (C). Discordancy and concordancy between biopsy and resection in relation to biopsy sample size; discordant TA: diagnosis in biopsy TC and in resection AC; concordant TT: biopsy and resection diagnosis TC; AA: biopsy and resection diagnosis AC (D). * p ≤ 0.05; ** p≤ 0.01 *** p ≤ 0.001.

Concordance in diagnosis between biopsy and resection was associated with increasing biopsy surface area, see Fig. 1D (discordant biopsies versus concordant biopsies for typical carcinoid (*p*=0.009) and atypical carcinoid (*p*=0.004)). Of 14 biopsies measuring <2 mm^2^, 11 cases (79%) were misclassified as TC. Likewise, 52% (15/29) of the discordant biopsies measured <4 mm^2^.

## Discussion

A small biopsy surface is associated with poor diagnostic accuracy in carcinoid classification. Biopsies taken during flexible bronchoscopy result in 59% of the patients misclassified as TC, compared to 30% of biopsies obtained with rigid bronchoscopy. This is, at least in part, explained by a difference in biopsy surface area on which the histological diagnosis is made, as an increase in cumulative biopsy surface to ≥4 mm^2^ leads to a significant reduction of the discordancy rate, and thus an increased in diagnostic accuracy.

Our data are in line with a recent retrospective study analyzing the accuracy of pre-operative biopsies for bronchial carcinoid tumors. The authors reported a 57% discrepancy when pre-operative biopsies were compared to postoperative pathology (Moonen et al., 2020). However, this study did not compare the influence of flexible with rigid biopsies and biopsy surface. Although discordant TC-AC diagnosis is not excluded in larger biopsies predominantly obtained via rigid biopsy, 52% of the discordant cases have a biopsy surface measuring <4 mm^2^. Conceptually, a preferred cumulative biopsy surface may be estimated that is associated with a higher diagnostic accuracy. We assume that a cumulative surface of 4 mm^2^ is equivalent to ± 4 bronchial biopsies of 1 mm^2^ tumor (Figure 2) or 2 biopsies of 2 mm^2^ tumor. Hypothetically, biopsies with a diameter of <4 mm^2^ can only be defined as bronchial carcinoid tumor, but not classified in TC or AC.

**Figure 2:**
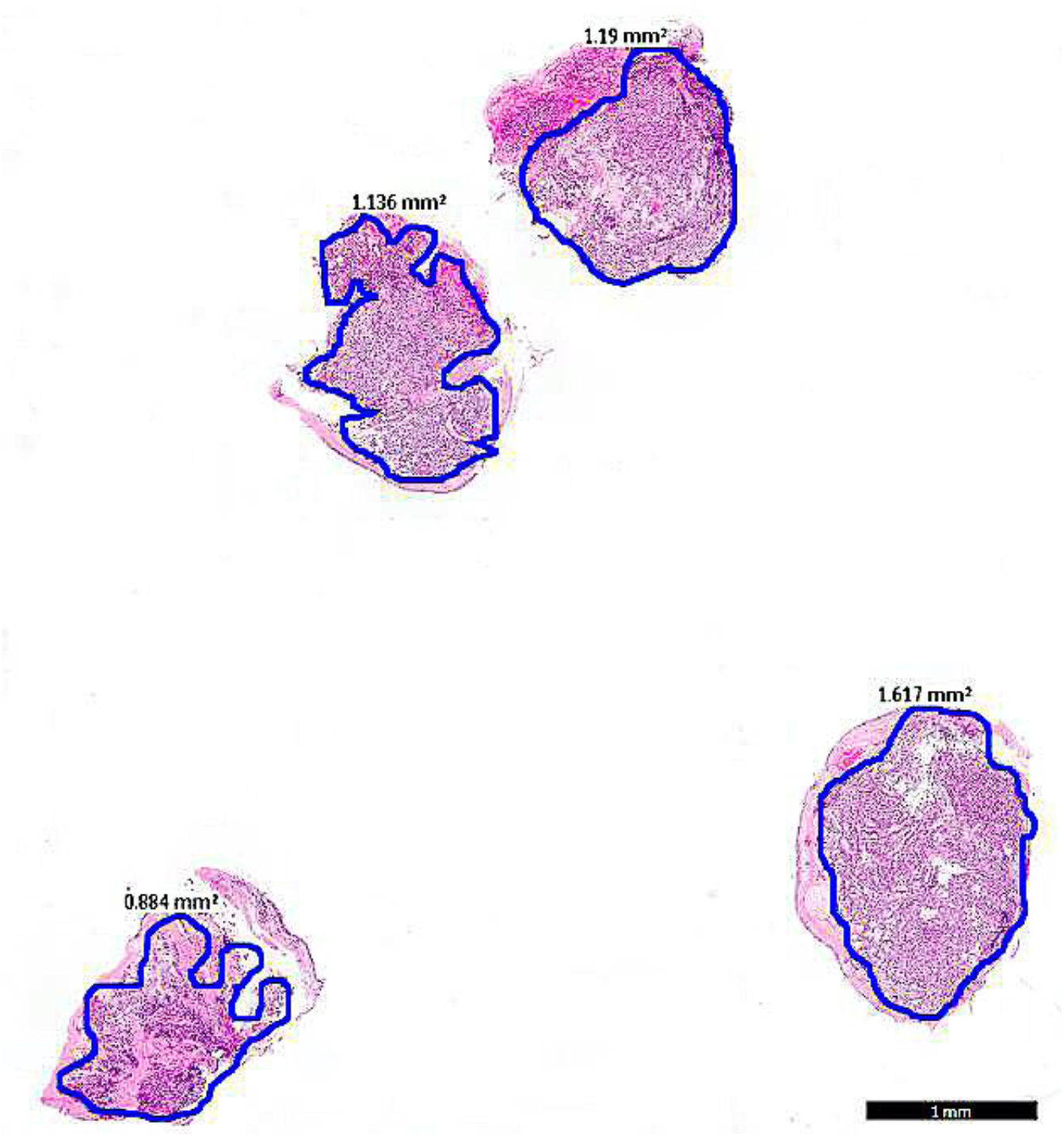
example of 4 flexible biopsies of 1 mm^2^ with bronchial carcinoid.

Flexible biopsies are easier to obtain and require less sedation compared to rigid biopsies. However, performing 4 biopsies in a relatively high vascularized tumor in non-anesthetized patients is challenging due to a risk of difficult to control bleeding. Preferably these biopsies should be performed in a controlled setting under general anesthesia via a rigid bronchoscopy. However, these procedures are predominantly performed in specialized centers but should be considered whenever clinically relevant: e.g. in patients with central bronchial carcinoid tumors suitable for curative endobronchial therapy, or patients unfit for surgery in whom bronchoscopic debulking could relieve symptoms of dyspnea or postobstructive pneumonia.

Thus, biopsy size does matter, which was previously shown in large cell neuroendocrine carcinoma, where neuroendocrine morphology was more frequently lacking in smaller biopsies (<5 mm) when compared to larger biopsies (Derks et al., 2019). In addition, for determination of PD-L1 in lung cancer, a biopsy size of <2 mm is associated with a 14% chance of false negative score (Bigras et al., 2018, Thunnissen et al., 2020). These examples, and findings from the current study, underscore the fact that small biopsy samples are associated with ‘false negatives’ /underdiagnoses.

A limitation of this study is that in our cohort, a slightly larger proportion of AC was observed than in the literature (E.M.B.P. Reuling, 2019). A possible explanation may be selection bias, as our center is a tertiary referral center for endobronchial treatment and more complex surgery. However, the higher proportion of AC allowed a more precise investigation of diagnostic accuracy for AC in the biopsies in a relatively small patient number.

## Conclusion

Histological classification in central carcinoid tumors is discordant in 45% of the biopsies, and diagnostic accuracy rises with larger biopsies. Classifying bronchial carcinoid in TC or AC in biopsies <4 mm^2^ should therefore be discouraged. A cumulative biopsy surface of at least 4 mm^2^ tumor is preferred to increase the diagnostic accuracy which helps in optimal treatment planning.

## Data Availability

The data that support the findings of this study are available on request from the corresponding author, [initials]. The data are not publicly available due to [restrictions e.g. their containing information that could compromise the privacy of research participants].

## Author contribution statement

ER, TR, ET and HD were responsible for the conception and design of the study and acquisition of data. ER and DN performed analysis and interpretation of the data. The article has been written by ER, TR, ET and critically revised by the authors who all gave approval for submission. Each author has participated sufficiently in the contributions of this article and agreed to be accountable for all aspects of the work in ensuring that questions related to the accuracy or integrity of any part of the work are appropriately investigated and resolved.

## Acknowledgement

The authors thank all of the involved clinicians, nurses, and technicians for dedicating their time and skills to this study.

